# Malaria Stratification Mapping in Thailand to Support Prevention of Re-establishment

**DOI:** 10.1101/2023.09.07.23295227

**Authors:** Donal Bisanzio, Prayuth Sudathip, Suravadee Kitchakarn, Jerdsuda Kanjanasuwan, Deyer Gopinath, Niparueradee Pinyajeerapat, David Sintasath, Jui A. Shah

**Affiliations:** Inform Asia: USAID’s Health Research Program, RTI International, Bangkok, Thailand; Division of Vector Borne Diseases, Department of Disease Control, Ministry of Public Health, Nonthaburi, Thailand; World Health Organization, Nonthaburi, Thailand; U.S. President’s Malaria Initiative, United States Agency for International Development (USAID), Regional Development Mission for Asia, Bangkok, Thailand

**Keywords:** stratification, prevention of re-establishment, localization, mosquito-borne disease, malaria elimination

## Abstract

Thailand aims to eliminate malaria by 2026, with 46 of the country’s 77 provinces already verified as malaria free. However, these provinces remain susceptible to the re-establishment of indigenous transmission that would threaten the national goal. Thus, the country is prioritizing national and subnational prevention of re-establishment (POR) planning while considering the spatial heterogeneity of the remaining malaria caseload. To support POR efforts, a novel non-modeling method produced a malaria stratification map at the tambon (subdistrict) level, incorporating malaria case data, environmental factors, and demographic data. The stratification analysis categorized 7,425 tambons into the following four risk strata: Local Transmission (2.9%), At Risk for Transmission (3.1%), High Risk for Reintroduction (2.9%), and Low Risk for Reintroduction (91.1%). The stratification map will support the national program to target malaria interventions in remaining hotspots and mitigate the risk of transmission in malaria-free areas.

---

Thailand aims to eliminate malaria by 2026. Over the past decade, malaria incidence in the total population has significantly decreased, declining from 3.6 cases per 1,000 people in 2012 to 0.2 cases per 1,000 people in 2021.^1^ This success in reducing malaria burden is due to intense intervention activities based on timely case investigation, case classification, and foci investigation.^2^ During 2016, Thailand formally shifted from a control to an elimination program by adopting the National Malaria Elimination Strategy (NMES) 2017–2026^3^ and the 1-3-7 surveillance approach^4^.

The World Health Organization (WHO) recommends that countries tailor interventions based on malaria stratification risk.^5^ This approach allows countries with low malaria incidence to promote targeted interventions in remaining hotspots of autochthonous transmission. Thailand’s Division of Vector-Borne Diseases (DVBD) has a subnational verification of elimination program, which has documented and celebrated that 46 of the country’s 77 provinces are malaria free. The program builds on the WHO’s rigorous criteria for national certification. However, even malaria-free provinces remain vulnerable to the reintroduction of malaria parasites and the re-establishment of indigenous transmission, threatening national progress toward elimination. Thus, subnational prevention of re-establishment (POR) planning is an essential component of a malaria elimination program^6^ to maintain the status of malaria-free areas while addressing remaining hotpots.

Malaria incidence in Thailand exhibits substantial heterogeneity, with malaria-free areas predominantly located in the central part of the country and remaining transmission areas along the borders with Cambodia, Myanmar, and Malaysia.^6^ To account for this heterogeneity, the DVBD implements malaria strategies at a focus level^7^, necessitating a stratification approach to support targeted POR interventions with increased resolution. This study presents a novel method for performing malaria stratification mapping in Thailand by using routine malaria surveillance data and environmental information to support POR planning.

The stratification followed the WHO’s guidelines^5^ and used data at the tambon (subdistrict) level from fiscal years (FYs) 2019 through 2022. A list of receptivity and vulnerability variables was compiled during discussions with the DVBD and partners. The selected variables included those recorded in Thailand’s malaria information system (MIS) and other contextual variables that could affect malaria transmission dynamics. The variables described the number of reported malaria cases population movement, demographic characteristics of reported cases, vector ecology, parasite characteristics, interventions, and environmental factors (a full list and data sources are reported in the Supporting Information). The stratification method accounted for the following national malaria foci classifications: active focus (A1), residual non-active focus (A2), cleared but receptive focus (B1), or cleared but not receptive focus (B2) (Table S1). One tambon would have several foci, with varying classifications.

The methodology aimed to create a stratification map based on foci classification and receptivity, demographic factors, and environmental characteristics to identify areas by risk of malaria re-establishment. The unsupervised approach was based on principal component analysis (PCA).^8^ The PCA estimates weights for each variable category based on its relationship with all other variables, rather than being chosen arbitrarily.^9, 10^ Investigating these correlations allows the PCA to condense multiple variables’ information into one score.^8^ This statistical method is most frequently used in economics to group people into wealth categories by using their assets.^11^

The PCA also included variable selection to identify only variables with high importance in classifying the tambons for malaria caseload and risk of re-establishment; variables also needed to cover all tambons within the study period (as described in the Supporting Information). In line with malaria stratification performed elsewhere^9, 10^, tambons were grouped into four strata. This grouping results in a manageable number of intervention packages for the DVBD yet maintains an informative level of detail (Figure S1).

The PCA’s grouping performance was assessed using the Kruskal-Wallis test for statistically significant differences of the selected variables among the strata.^12^ All statistical analyses were performed using the R programming language.^13^ The stratification process was provided to the DVBD as an ad hoc statistical software created using Jamovi (www.jamovi.org), which is open-source statistical software.

Among Thailand’s 7,425 tambons, the stratification analysis assigned 216 (2.9%), 228 (3.1%), 216 (2.9%), and 6,765 (91.1%) tambons to Stratum 1, Stratum 2, Stratum 3, and Stratum 4, respectively. The results of the PCA showed good grouping performance, with the Kruskal-Wallis test showing that the values of the variables used were significantly different across strata (p<0.05).

Given the characteristics of the tambons in each stratum, the following labels were assigned to represent each stratum’s probability to report autochthonous cases:

- Stratum 1: Local Transmission
- Stratum 2: At Risk for Transmission
- Stratum 3: High Risk for Reintroduction
- Stratum 4: Low Risk for Reintroduction.

Stratum 1: Local Transmission included tambons with the highest malaria caseload, with a median (MD) incidence equal to 2.7 malaria cases per 1,000 people (interquartile range [IQR]: 0.23–2.4 cases per 1,000 people) (Table 1). Tambons in Stratum 4: Low Risk for Reintroduction had the lowest malaria incidence from FY 2019 to FY 2022 among all the strata (MD 0; IQR: 0–0). Tambons allocated in Stratum 2: At Risk for Transmission and Stratum 3: High Risk for Reintroduction had a median malaria incidence below the elimination threshold of 1 case per 1,000 people (Table 1).

**Table 1.** Characteristics of the tambons of each stratum created using the PCA approach. The stratum population was equal to the population of tambons of the stratum. The estimated number of households was obtained by dividing the tambon population by the mean number of people per household (2.7 people per household). Table 1 is based on data recorded from 2019 through 2022.

| Characteristics | Stratum 1:<br>Local<br>Transmission | Stratum 2:<br>At Risk for<br>Transmission | Stratum 3:<br>High Risk for<br>Reintroduction | Stratum 4:<br>Low Risk for<br>Reintroduction |
| --- | --- | --- | --- | --- |
| Tambons | 216 | 228 | 216 | 6,765 |
| Population<br>(proportion) | 761,739<br>(1.1%) | 830,078<br>(1.2%) | 1,174,153<br>(1.6%) | 66,835,383<br>(96.1%) |
| Estimated number of<br>households | 282,125 | 307,436 | 434,871 | 24,753,845 |
| PCA variables<br>Median (IQR) |  |  |  |  |
| Mean annual<br>incidence<br>(per 1,000 people) | 2.73<br>(0.23; 2.4) | 0.17<br>(0.08; 0.19) | 0.05<br>(0.04; 0.06) | 0<br>(0; 0) |
| Mean percentage (%)<br>of A1 foci per year | 12.54<br>(3.85; 16.07) | 0.72<br>(0; 1.25) | 0<br>(0; 0) | 0<br>(0; 0) |
| Years since last<br>reported A1 foci | 1<br>(0; 1) | 3<br>(3; 4) | 4<br>(4; 4) | 4<br>(4; 4) |
| Mean percentage (%)<br>of *Plasmodium*<br>*falciparum* cases | 4.29<br>(0; 5) | 0<br>(0; 0) | 0<br>(0; 0) | 0<br>(0; 0) |
| Mean percentage (%)<br>of indigenous cases | 30.24<br>(0; 63.83) | 0<br>(0; 0) | 0<br>(0; 0) | 0<br>(0; 0) |
| Mean percentage (%)<br>of plantation worker<br>cases | 8.23<br>(0; 10.42) | 0<br>(0; 0) | 0<br>(0; 0) | 0<br>(0; 0) |
| Mean percentage (%)<br>of Thai cases | 40.55<br>(0; 91.35) | 0<br>(0; 0) | 0<br>(0; 0) | 0<br>(0; 0) |
| Mean percentage (%)<br>of cases with a travel<br>history outside<br>Thailand | 18.75<br>(0; 32.79) | 4.86<br>(0; 0) | 0<br>(0; 0) | 0<br>(0; 0) |

Tambons in Stratum 1: Local Transmission showed a higher mean percentage of A1 foci (MD 12.5%) compared with the other strata, in which the mean percentage of A1 foci was less than 1% (Table 1). Stratum 1: Local Transmission also showed recent reporting of A1 foci, whereas most of the tambons in the other strata have not reported autochthonous transmission in at least 3 years (Table 1). The fraction of cases caused by *Plasmodium falciparum* was higher in the tambons belonging to Stratum 1: Local Transmission compared with those in the other strata (Table 1). Thai citizens and plantation workers represented more reported cases in Stratum 1: Local Transmission compared with other strata (Table 1).

The spatial pattern of the strata showed that Stratum 1: Local Transmission is mostly aggregated at the borders with Myanmar, Malaysia, and Cambodia (Figure 1). The analyses also identified the tambons in low endemic provinces classified as Stratum 1: Local Transmission and Stratum 2: At Risk for Transmission (Figure 1). People living in Strata 1, 2, and 3 represented 1.1%, 1.2%, and 1.6% of Thailand’s population, respectively (Table 1). Accounting for the mean number of household inhabitants, 282,125 households are located in the tambons assigned to Stratum 1: Local Transmission.

**Figure 1.**
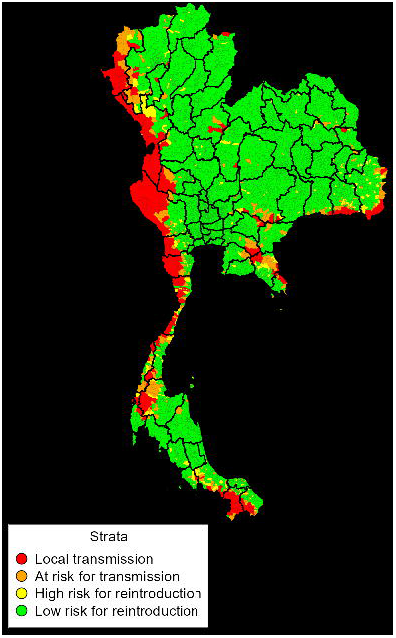
Malaria stratification map for Thailand using data from 2019 through 2022

The stratification results captured the known high heterogeneity of local transmission risk, but at the tambon level to better support POR decision-making and interventions. The method showed good grouping performance to identify areas in which the tambons are still reporting autochthonous cases. Furthermore, the results highlight additional areas at high risk of malaria transmission despite not having recently reported autochthonous cases; this novel information will support meaningful POR planning.

The stratification analysis showed that all tambons that still have A1 foci belonged to Stratum 1: Local Transmission (Table 2). A high fraction of foci in these tambons are classified as residual non-active (A2) and because of the frequent transition of A2 foci to A1 classification^14^, there is high risk for local transmission to resume within tambons in Strata 1 and 2. These strata were mostly identified along international borders, following well-known patterns in the Greater Mekong Subregion.^7^ High human mobility across these borders, a suitable environment, and limited access to must-reach populations continue to drive malaria transmission in these tambons.^4, 15, 16^ To reduce the risk of outbreaks by imported cases in areas close to the western border, the DVBD implements malaria interventions tailored for mobile and migrant populations.^17^ However, the persistence of transmission in border areas indicates a need for new strategies to interrupt transmission.^18^

**Table 2.**
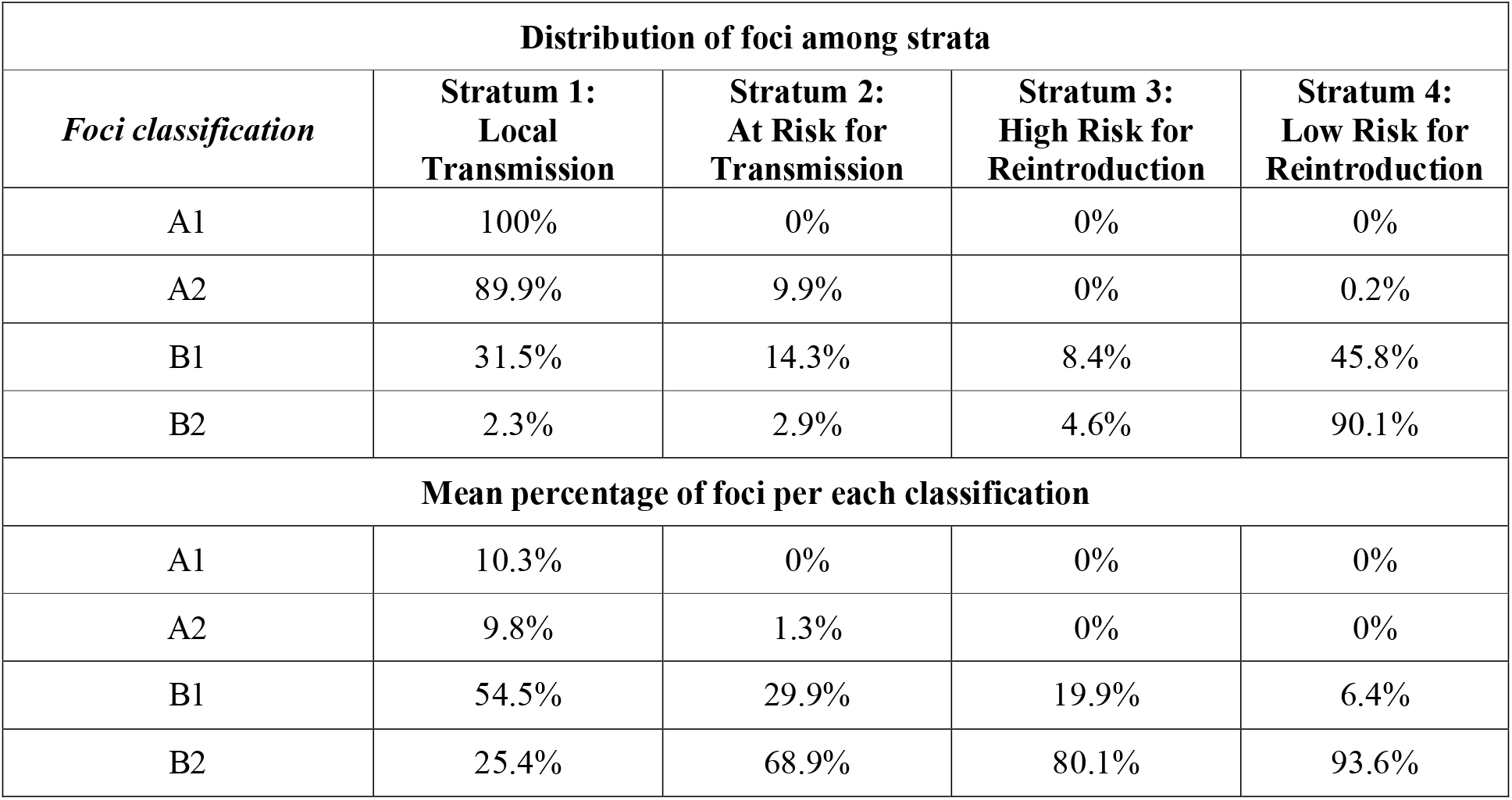
Distribution of 2022 malaria foci per malaria stratum.

The results showed that 97.7% of Thailand’s population resides in Stratum 3: High Risk for Reintroduction and Stratum 4: Low Risk for Reintroduction. In these strata, local transmission has not been reported for more than 3 years and the DVBD is launching POR plans. However, tambons in Stratum 3: High Risk for Reintroduction have a higher fraction of cleared but receptive foci (B1, 19.9%) compared with those in Stratum 4: Low Risk for Reintroduction (6.4%). The presence of receptive foci in these malaria-free areas requires tailored POR planning, based on risk and available resources, to maintain Thailand’s current successes and to accelerate toward malaria elimination.

This study also aimed to build a stratification approach that could be easily adopted and maintained by the country as part of routine surveillance and strategic planning at both national and subnational levels. Compared with stratification approaches used elsewhere^19, 20^, this approach has two substantial benefits. First, the approach highlights Thailand’s use of routine surveillance data, which supports country leadership and reduces reliance on imputed data. Second, the DVBD can learn the approach and maintain it with annual updates, as the epidemiology warrants. The stratification analysis and results were programmed into Jamovi and accompanied by sufficient training to support sustained use in the country through the malaria elimination goal.

## Data Availability

All data produced in the present study are available upon reasonable request to the authors

## Supporting information

Data Dictionary and Data Selection, Data Collection, Stratification Methodology, Variable Selection, Jamovi Module

Table S1. Classification of malaria foci in Thailand 12

**Figure S1.**
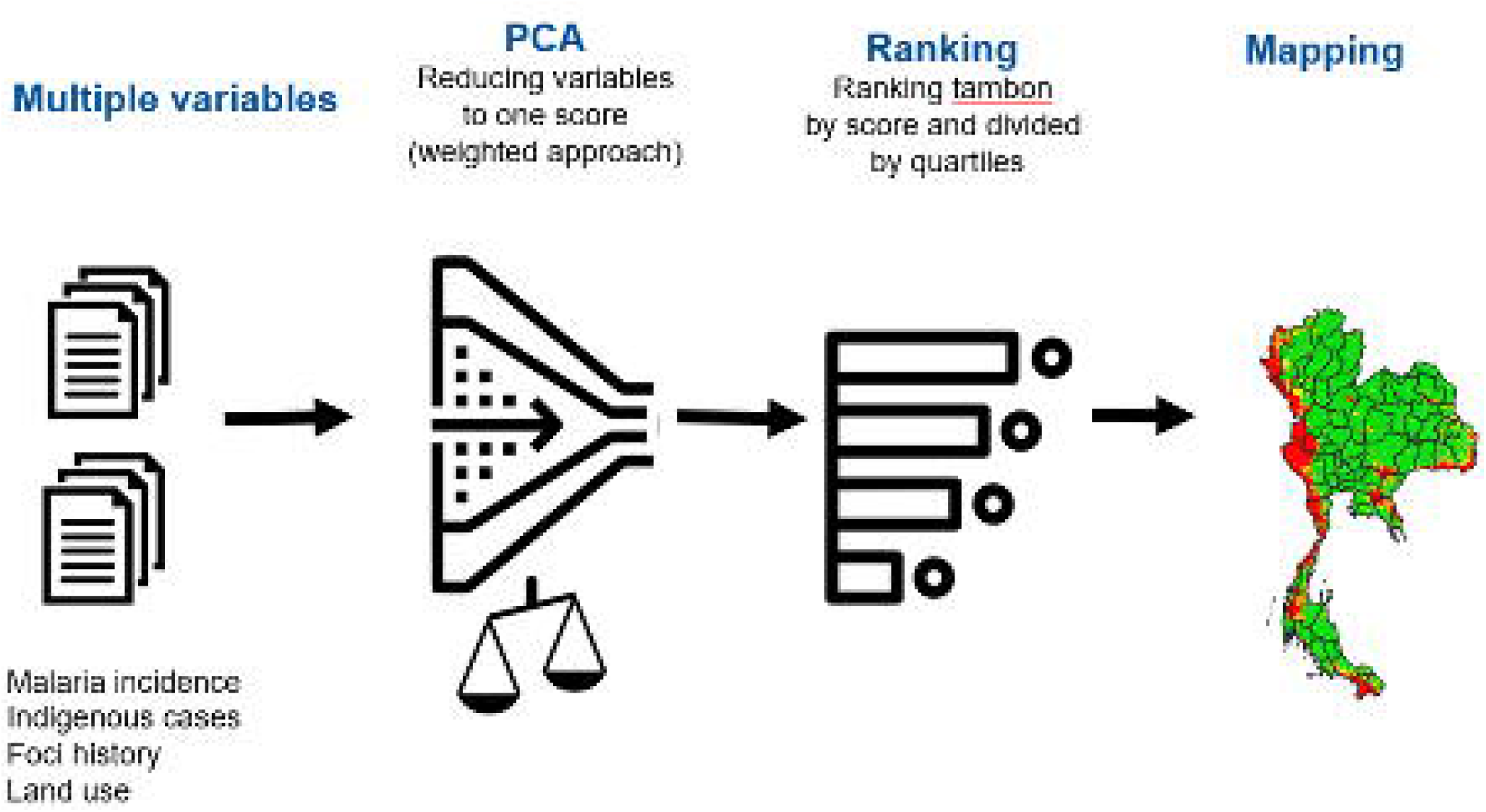
Flow chart of Thailand’s malaria stratification mapping approach

**Figure S2.**
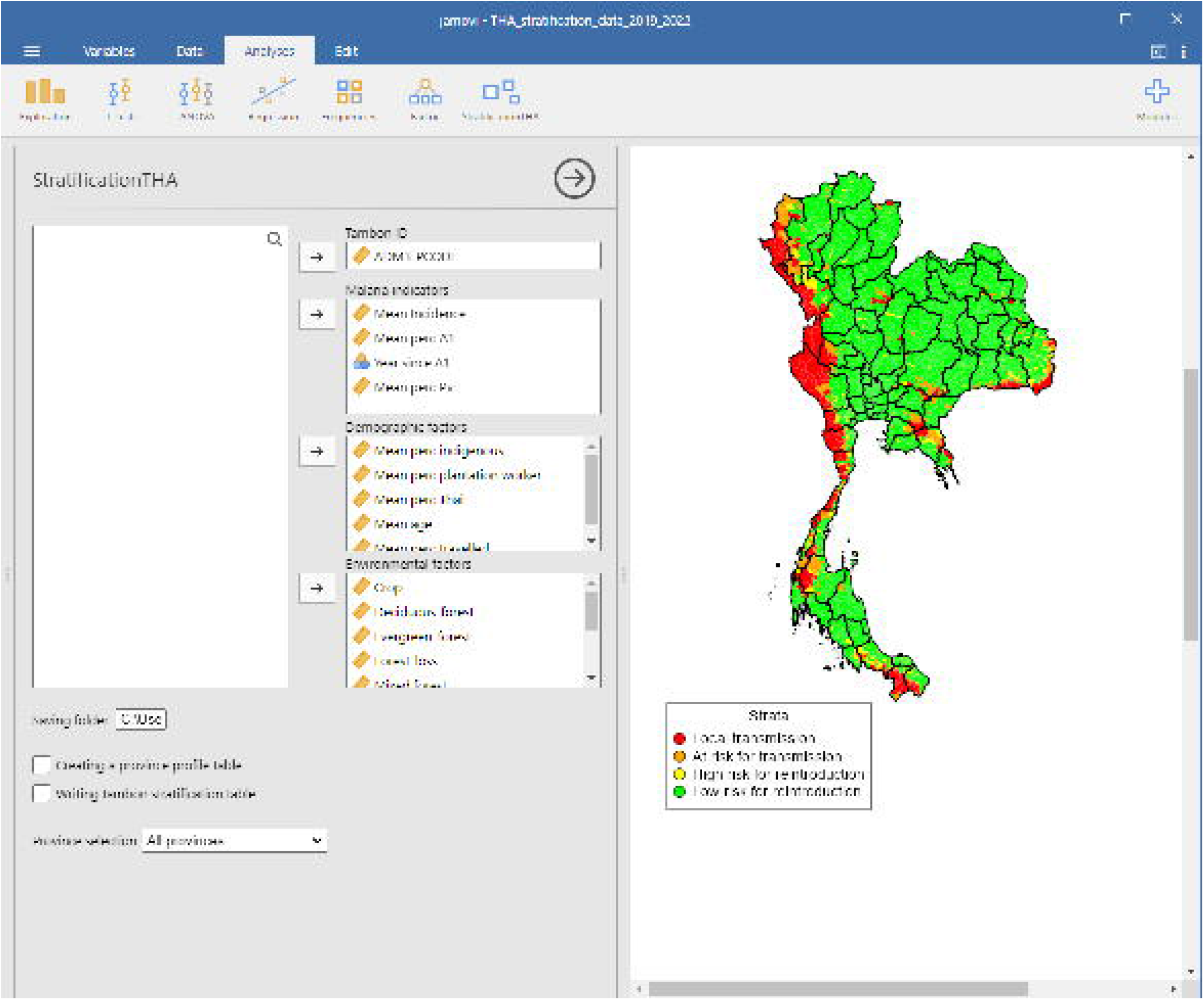
Screenshot of Thailand’s malaria stratification module installed in the Jamovi software

## Funding

This study was made possible by the generous support of the American people through the U.S. President’s Malaria Initiative (PMI) and United States Agency for International Development (USAID), under the terms of Cooperative Agreement AID-486-LA-15-00002 for Inform Asia: USAID’s Health Research Program.

## Disclaimer

The contents of this article are the responsibility of the authors and do not necessarily represent the official position of PMI, USAID, or the U.S. Government. DG is a staff member of the World Health Organization (WHO) and is responsible for the views expressed in this publication, which do not necessarily reflect the decisions or policies of the WHO.

## Notes

### Competing Interest Statement

The authors have declared no competing interest.

