## Supplementary material for "Malaria Stratification Mapping in Thailand to Support Prevention of Re-establishment": Supplemt information

**Supporting Information**

*Data Dictionary and Data Selection*

The stratification approach for Thailand’s prevention of re-establishment (POR) planning was based on the World Health Organization’s stratification guidelines and other best practices.^1, 2, 3, 4, 5^. We compiled a list of potential variables for consideration in the development of malaria stratification maps for Thailand during discussions with the Division of Vector-Borne Diseases (DVBD) and partners. The list of proposed variables related to receptivity and vulnerability included malaria indicators recorded in Thailand’s malaria information system (MIS) and other contextual variables that could affect malaria transmission dynamics in Thailand, including the following:

- Malaria burden: Malaria incidence, foci classification, parasite species, and source of infection (autochthonous or imported cases)
- People movement: Travel history outside Thailand
- Demographic data: Age structure, gender ratio, socio-economic status, and fraction of population per profession group
- Environmental factors: Elevation and percentage of territory cover by different habitat type (e.g., crop, forest, urban)
- Vector data: Species distributions, mosquito abundance, feeding and resting behaviors, and insecticide resistance
- Malaria interventions: Reactive case detection (RACD), proactive case detection (PACD), indoor residual spraying (IRS), and social and behavior change (SBC) activities
- Parasite characteristics: Parasite species and parasite genomics.

This aspirational list of variables was then reviewed for spatio-temporal coverage and eligibility, as described in the Variable Selection section of this document.

*Data Collection*

Routine malaria surveillance data were extracted from Thailand’s robust national MIS. The stratification was performed by using data at the tambon (subdistrict) level from fiscal years (FYs) 2019 through 2022. The study utilized fiscal years (October to September) because Thailand’s malaria program and database are based on fiscal year targets.

For environmental data, we downloaded land cover rasters with a 100-meter (m) resolution from the Copernicus website (<https://land.copernicus.eu/global/products/lc>).^6^ Using the rasters, we calculated the percentage of tambon territory covered by crops, mixed forest, evergreen broad leaf forest, deciduous forest, built areas, and rice fields. We also included information about deforestation because it has been associated with malaria transmission in Thailand.^7^ Latest available raster data about forest disturbance on a global scale from 2015 to 2019 were obtained from the Global Forest Change website (<http://earthenginepartners.appspot.com/science-2013-global-forest>).^8^ The values reported in the data create a disturbance index indicating the level of disturbance on a scale from 0 (no disturbance) to 17 (highest level of disturbance) at a 25-m resolution. We calculated the mean annual disturbance index for each tambon.

*Stratification Methodology*

In line with malaria stratification efforts performed elsewhere^2, 9^, tambons were grouped into four strata based on foci classification and receptivity, demographic factors, and environmental characteristics (and, thus, risk for re-establishment). We used principal component analysis (PCA) technique, which condenses multiple categorical variables’ information in a score by investigating their correlations.^10^ This unsupervised approach based on a variance analysis allows handling of variables with high correlation.^10^

The score of each tambon was calculated by summing the PCA weight values of the tambon’s variable categories. The tambons were divided into four based on score quartiles (Figure S1). Each quartile represented one of the four strata: Stratum 1 included tambons with the highest malaria burden and Stratum 4 comprised tambons with the lowest malaria burden. The PCA’s performance was assessed with the Kruskal-Wallis test^11^ for significant differences in malaria indicators among the strata.

*Variable Selection*

Variables were evaluated for inclusion in the stratification analysis; the PCA only used variables meeting the following criteria:

1. Spatial resolution at the tambon level
2. Available for all tambons without missing data
3. Data values in the selected time window (FYs 2019 to 2022)
4. Higher percentage of explained variance than the average explained variance if all variables were equal (100% ÷ number of PCA variables).

These variables address both vulnerability, or “importation risk” based on the human side of malaria epidemiology, and receptivity, covering the vector and environment sides.

The PCA provides a score for each tambon by summing its variable weights, so the method cannot perform imputation of missing data. Using data that are available for only a few tambon would introduce a bias in the score calculation because of missing weights in those tambons with missing values in the dataset. We decided not to apply available imputation methods due to their low performance that would introduce an error noise into the analysis.

Among the initially listed variables, the following variables fulfilled the first three inclusion criteria, and therefore were included in the stratification analysis to test whether they also fulfilled the fourth criterion:

Malaria variables

- Malaria incidence
- Historical foci classification (up to 4 years)
- Percentage of cases infected by *Plasmodium vivax*
- Percentage indigenous cases.

Environmental variables

- Percentage of land occupied by tropical forest
- Percentage of land occupied by mixed forest
- Percentage of land occupied by tree plantations
- Percentage of land occupied by croplands
- Percentage of land occupied by built area
- Percentage of land occupied by seasonal water bodies
- Percentage of land occupied by permanent water bodies
- Level of human forest disturbance.

Demographic variables

- Case age
- Case sex
- Case travel history
- Case residential status
- Case occupation.

The variable list does not include entomological data and information about resistance in malaria parasite because they are not available for the entire country. Intervention data were not included in the stratification analysis to allow using the stratification classification in analyses assessing intervention efficacy. Using strata created by including interventions in modeling assessing intervention efficacy would violate the assumption that model variables are independent and thus prevent use in future effectiveness analyses.

We performed the PCA by using all of the variables that fulfilled the first three inclusion criteria and calculated the percentage of variance explained by each of them. All of the variables that explained more than the average explained variance (100% ÷ number of variables) were included in the PCA used to perform the malaria stratification.

The selected variables for the stratification PCA are as follows:

- Mean tambon annual incidence
- Mean percentage of foci classified as active focus (A1) per year
- Year since an A1 foci has been reported in the tambon
- Mean of the annual percentage of *Plasmodium falciparum* cases among reported cases in the tambon
- Mean of the annual percentage of autochthonous cases among reported cases in the tambon
- Mean of the annual percentage of plantation workers among reported cases in the tambon
- Mean of the annual percentage of reported cases with a travel history outside Thailand in the tambon
- Mean of the annual percentage of Thai citizens among reported cases in the tambon.

*Jamovi Module*

A focus of the analyses presented in this manuscript was to build in-country capacity to perform malaria stratification for Thailand. Because no off-the-shelf stratification software can perform this stratification, we worked with the DVBD to develop an easy-to-use tool to facilitate the continued use of the stratification analysis method and leadership in using the results.

We assessed different software solutions and selected the Jamovi software to develop the stratification tool because—for long-term sustainability—we needed a free, open-source software with high flexibility. Jamovi allows performing statistical analysis using the R language but with a simple graphic user interface (Figure S2) and customizable, easily installed modules of analytical tools. We created a stratification module that can be installed in Jamovi once the user provides the dataset for the analysis. The module then produces tables and maps showing the results of the stratification for all of Thailand’s tambons, grouped by province. The resulting stratification map can be used in risk planning because it highlights the areas at high risk of reporting indigenous cases.

**Table S1. Classification of malaria foci in Thailand ^12^**

| **Focus classification** | **Definition** |
| --- | --- |
| A1 | Active focus: Reported indigenous transmission in the current year |
| A2 | Residual non-active focus: No indigenous cases in the current year but with indigenous cases during the previous 3 years |
| B1 | Cleared but receptive focus: No indigenous transmission in at least 3 years but suitable environment for vector *Anopheles spp.* mosquitoes |
| B2 | Cleared but receptive focus: No indigenous transmission in at least 3 years but unsuitable environment for vector *Anopheles spp.* mosquitoes |
